# Causal associations between niacin bluntness and schizophrenia: a GWAS and Mendelian randomization study

**DOI:** 10.1101/2024.11.24.24317680

**Authors:** Liya Sun, Fangyu Chen, Mo Li, Changchang Liu, Jie Jiang, Yuanyuan Dai, Jinfeng Wang, Yan Gao, Lin He, Shengying Qin, Chunling Wan

**Affiliations:** Shanghai Mental Health Center, School of Medicine, Shanghai Jiao Tong University, Shanghai, 200030, China; WuHu Hospital of Beijing Anding Hospital, Capital Medical University (WuHu Fourth People’s Hospital), Wuhu, 241003, China; Department of Cardiology of The Second Affiliated Hospital, School of Medicine, Zhejiang University, Hangzhou 310009, China; Shandong Mental Health Center, Jinan, 250014, China; Bio-X Institutes, Key Laboratory for the Genetics of Developmental and Neuropsychiatric Disorders (Ministry of Education), Shanghai Key Laboratory of Psychotic Disorders, Shanghai Mental Health Centre, Shanghai Jiao Tong University, Shanghai, 200030, China

## Abstract

**Background and Hypothesis:** Niacin skin bluntness is a promising biomarker for schizophrenia, especially in precision medicine, as it helps identify a distinct subgroup (around 1/3) of patients with schizophrenia who experience severe functional impairment. However, research has not clarified if this phenotype is just a byproduct of the onset of the disease or is involved in the etiology of schizophrenia. We hypothesize that niacin bluntness reflects causal alterations in schizophrenia.

**Study Design:** We firstly conducted a quasi-genome-wide association analysis within schizophrenia patients to identify instrumental SNPs for niacin response. Then a two-sample bi-directional Mendelian randomization (MR) analysis was implemented to estimate the potential causal effects between niacin response and schizophrenia.

**Study Results:** 25 independent SNPs showed a trend of genome-wide significant association (p<E-05) with niacin response in schizophrenia. In the MR analysis, the F statistics of the two instrumental SNPs for niacin response is 30.77, indicating a proper strength. As heterogeneity of the SNPs was detected (p<0.05), the result of inverse variance weighted method for multiplicative random effects was adopted for evaluation of the detected causal effect (OR=1.272, p=9.46E-03). In reverse MR analysis, none of the methods supported a causal effect of schizophrenia on niacin response (all p>0.05).

**Conclusions:** Our results revealed that the attenuated niacin response caused schizophrenia but not vice versa. Etiological studies on niacin bluntness in schizophrenia are needed and will pave the way for biomarker-guided personalized treatment in future.

## 1. Introduction

Schizophrenia is a severe mental disorder resulting from complex interactions between genetic and environmental factors. Approximately one-third of patients with schizophrenia are resistant to current treatments ^1^. Historically, etiological research on schizophrenia has been undermined by the subjective nature of existing diagnostic criteria and heterogeneity among patients in terms of both symptomatic manifestations and pathogenesis. Searching for the objective markers of schizophrenia could assist with diagnosis and facilitate precise management. Niacin bluntness, a unique skin feature manifested in the niacin flushing test, is a promising biomarker of schizophrenia that was first proposed by Horribin in 1980^2^ and has been repeatedly confirmed in various cohorts with high specificity^3^. Niacin flushing status can be semi-quantitatively evaluated using flushing scores ^4^. The proportion of patients with schizophrenia with typical niacin bluntness (i.e., having niacin flushing scores below the lowest boundary of the normal range) is estimated to be within 25%–33%, forming a distinctive biological subtype of schizophrenia ^5^. Patients with niacin bluntness were found to have an increased prevalence of negative symptoms and severe functional impairments^6,7^. However, existing research has not clarified whether niacin bluntness is a phenotypic result of schizophrenia or whether it reflects certain causative factors contributing to disease onset. If the latter is true, this phenotype would be much more clinically meaningful. Further research should be conducted to delineate the molecular basis of niacin bluntness to provide potential drug targets for precise and efficient treatment for individuals with schizophrenia in the niacin-blunted subgroup.

In this study, we evaluated the causative effect of niacin bluntness on schizophrenia by conducting a preliminary genome-wide association analysis (GWAS) in a sample of schizophrenia patients to identify potential instrumental single nucleotide polymorphisms (SNPs) associated with niacin bluntness. Two-directional Mendelian randomization (MR) analysis was conducted to estimate the potential causative relationships between niacin flushing status and schizophrenia, utilizing the largest GWAS data on schizophrenia in East Asia ^8^.

## 2. Methods

### 2.1 GWAS of the niacin bluntness phenotype in participants with schizophrenia

#### 2.1.1 Sample recruitment

Participants with schizophrenia were recruited from inpatients at the Fourth People’s Hospital of Wuhu, Anhui Province, China. The inclusion criteria were: (1) having a diagnosis of schizophrenia according to ICD-10 (International Classification of Diseases, 10th Revision) and (2) being between 15–75 years of age. The exclusion criteria were: (1) a diagnosed neurological disease, such as epilepsy, cerebral tumor, or severe head injury; (2) the use of nonsteroidal or steroidal anti-inflammatory drugs within the previous 14 days; (3) severe skin diseases or diseases that would induce significant immune responses, such as lupus and asthma; and (4) pregnancy. This study was approved by local institutional ethics committee.

#### 2.1.2 Niacin flushing test and scoring

Niacin skin flushing tests were performed according to previously described methods with modifications to the concentrations of aqueous methyl nicotinate (AMN). In this study, we used three AMN concentrations (0.001M, 0.01M and 0.1M). In brief, the participants’ forearms were administered AMN patches of different concentrations for 1 min, then photographed every 5 min for 20 min, resulting in four photos per participant. The flush responses were scored using software developed in-house and rechecked by an experienced researcher on a 4-point scale (0 = *no erythema*, 1 = *incomplete erythema*, 2 = *complete erythema matching the area of the patch*, and 3 = *erythema beyond the patch area*). The sum of the 12 flushing scores (3 AMN concentrations × 4 time points) was defined as the total niacin response score for each participant.

#### 2.1.3 DNA extraction and storage

Peripheral venous blood samples were collected with EDTA anticoagulant tubes. Genomic DNA was extracted by standard procedures using an AxyPrep Blood Genomic DNA Miniprep Kit (Axygen, USA). All extracted DNA samples were preserved at -80°C before subsequent experiment.

#### 2.1.4 ASA chip experiment and data preprocessing

Single nucleotide polymorphism (SNP) genotyping was performed using an Illumina Asian Screening Array (ASA)-24 v1.0 BeadChip, following the Illumina Infinium HTS Assay protocol (Illumina, San Diego, CA, USA). The array platform contained 659,184 SNPs polymorphisms across the genome. Standard quality control (QC) filters were applied to exclude samples with a missing genotype rate >5% or based on a high heterozygosity rate (>3 × sigma). Duplicates and related individuals were removed using IBD analysis (PI-HAT ≥ 0.1875). SNPs were excluded because of a missing genotype rate >5%, being inequilibrium in the Hardy-Weinberg equilibrium test (*p* < 1E-06), or minor allele frequency (MAF) of the SNP <1%. After QC filtering, 464 samples and 488,130 SNPs were retained for subsequent SNP imputation. The imputation process was mapped to the ChinaMAP phase v1 reference panel using an online server (http://www.mbiobank.com/imputation/), and only high-quality SNPs (*R*^2^ > 0.3) were retained for further SNPs QC, as mentioned above. Finally, 6,777,277 SNPs were used for the subsequent association analysis. In addition, the population substructure was evaluated using principal component analysis (PCA) with the top five components being used as covariates in the association analysis to correct for population stratification.

#### 2.1.5 Genome-wide association analysis and post-GWAS bioinformatic analyses

The association between single SNPs and the overall niacin response score was tested by linear regression using PLINK, after correcting for the top five principal component scores, age, and sex. Quantile-quantile plots and Manhattan plots were generated using the qqman R package. Regional plots were generated using LocusZoom software (https://locuszoom.sph.umich.edu). We conducted annotation, functional mapping, gene-based analysis, and gene-set analysis of the results of the GWAS of niacin responses using the SNP2GENE function module of the online software Functional Mapping and Annotation (FUMA, https://fuma.ctglab.nl).

### 2.2 Mendelian randomization analysis

#### 2.2.1 Study design and data sources

The causal relationship between niacin response and schizophrenia was estimated using a two-sample Mendelian randomization (MR) analysis. Mendelian randomization (MR) is a method of studying the causal effects of modifiable exposures (i.e., potential risk factors) on health, social, and economic outcomes using genetic variants associated with the specific exposures of interest^9^. The analysis involves a risk factor (exposure) and a phenotype (outcome), consisting of three steps: 1) collecting GWAS summary-level data for the exposure, 2) exploring genetic variants from the GWAS summary data to serve as instrumental variables, and 3) estimating the causal effects of the exposure on the outcome using instrumental variables. We performed a bidirectional MR analysis. For niacin response, the associated SNPs below a threshold of *p* < .000001 in the previous GWAS were selected as candidate instrumental variables. For schizophrenia, summary-level data from a previously published GWAS^8^ in East Asians were used, involving 22,778 cases and 35,362 controls. The associated SNPs below a threshold of *p* < 5*10E-8 were obtained. A linkage disequilibrium threshold of *r*^2^ < .001 was used, based on the 1000 Genomes Project East Asian data, to identify independent SNPs.

#### 2.2.2 Mendelian randomization estimates

The R package TwoSampleMR was used to implement bidirectional MR estimates. Two SNPs were filtered out and analyzed as instrumental variables for niacin response, and 19 SNPs were analyzed as instrumental variables for schizophrenia (Supplementary Table 1). When the niacin response was taken as the exposure, heterogeneity was detected in the data structure, and we applied the inverse variance weighted (multiplicative random effects) method for MR estimation. When the schizophrenia phenotype was considered as the exposure, no heterogeneity was detected, thus we applied classical MR methods (e.g., MR-Egger and inverse variance weighted). Genetic pleiotropic assessment, single SNP effect analysis, and sensitivity analysis (leave-one-out method) were also conducted.

## 3. Results

### 3.1 GWAS results and functional analysis

We recruited 473 patients with schizophrenia for this study. The demographic characteristics of the participants and rough distribution of the niacin flushing scores are shown in Table 1. About one third of the patients’ niacin total flushing scores were under 10, while only 66 patients had total flushing scores above 20. No association of sex or age with the total flushing score were revealed. The GWAS of the niacin response scores revealed that two top SNPs (rs2993439, upstream of the *LGR6* gene on Chromosome 1, *p* = 8.32E-08, β=3.066; rs3010092, in the intron of *LRG6*, p=8.32E-08, β=3.066) had a trend of genome-wide significant association (*p* < 5E-08) with the total niacin flushing score in patients with schizophrenia. The Manhattan, quantile-quantile, and LocusZoom plots are shown in Figure 1A–C. Supplementary Table 1 includes additional information on the SNPs that reached the suggested threshold (*p* < 1E-05). Twenty-five suggestive significant independent SNPs were identified via the FUMA platform (Supplementary Table 2). A total of 122 potential genes involved in the niacin response were identified by positional mapping and chromatin interaction mapping of FUMA (Supplementary Table 3). Although none of the genes reached the genome-wide threshold of significance, MAGMA Tissue Expression Analysis revealed a trend of enrichment of genes highly expressed in the brain, for example, the anterior cingulate cortex and amygdala (all *p* <.10; Supplementary Figure).

**Table 1.** Characteristics of the schizophrenia patients in GWAS for niacin response.

| Items | Niacin flushing total score | | | | F/$\chi^2$ statistic | p value |
| --- | --- | --- | --- | --- | --- | --- |
|  | Overall | 0 to 10 | 11 to 20 | 21 to 36 |  |  |
| Number of patients | 473 | 135 | 272 | 66 |  |  |
| Gender (male, n(%)) | 280 (60.09) | 88 (65.19) | 152 (57.14) | 40 (61.54) | 2.48 | 0.289 |
| Age (years, mean (SD)) | 47.00 (12.12) | 48.01 (12.97) | 46.75 (11.90) | 45.84 (11.27) | 0.81 | 0.446 |
| Age of on set (years, mean (SD)) | 27.89 (9.97) | 28.01 (9.94) | 27.63 (10.18) | 28.80 (9.23) | 0.37 | 0.693 |
| Education (n(%)) |  |  |  |  | 4.83 | 0.566 |
| illiteracy | 46 (10.29) | 17 (13.08) | 23 (9.06) | 6 (9.52) |  |  |
| primary school | 95 (21.25) | 26 (20.00) | 54 (21.26) | 15 (23.81) |  |  |
| middle/high school | 282 (63.09) | 80 (61.54) | 166 (65.35) | 36 (57.14) |  |  |
| college/university | 24 (5.37) | 7 (5.38) | 11 (4.33) | 6 (9.52) |  |  |
| Marriage (n(%)) |  |  |  |  | 11.67 | 0.070 |
| married | 102 (22.17) | 24 (18.05) | 64 (24.43) | 14 (21.54) |  |  |
| divorced | 78 (16.96) | 18 (13.53) | 41 (15.65) | 19 (29.23) |  |  |
| widowed | 25 (5.43) | 9 (6.77) | 14 (5.34) | 2 (3.08) |  |  |
| unmarried | 255 (55.43) | 82 (61.65) | 143 (54.58) | 30 (46.15) |  |  |

**Figure 1.**
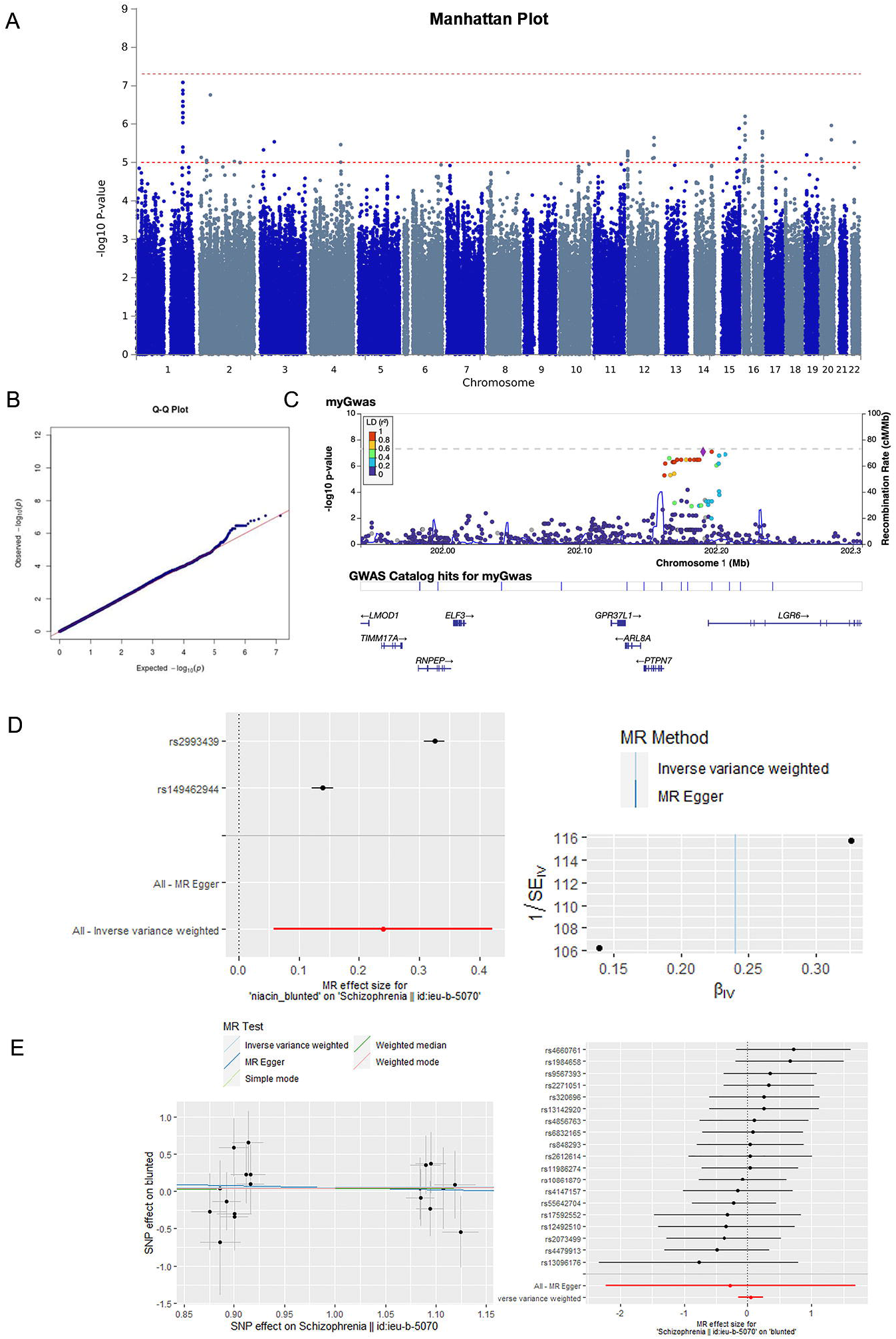
Genome-wide association analysis results of niacin skin response in schizophrenia and two directional Mendelian randomization (MR) results between niacin response and schizophrenia. For the Genome-wide association analysis (GWAS) results, Manhattan (Figure 1A), quantile-quantile (Figure 1B), and LocusZoom plots (Figure 1C) for the most significant sites are shown. For the MR results, forest plots and funnel plots of single nucleotide polymorphisms (SNP) effect estimates of the causal effect of niacin response on schizophrenia are shown in Figure 1D, and scatter plots and forest plots of single SNP effects of the causal effect of schizophrenia on niacin response are shown in Figure 1E.

### 3.2 The causal effect of niacin response on schizophrenia

The potential causal effect of niacin response on schizophrenia was explored. Two SNPs were filtered out by software as instrumental variables for the niacin response (Supplementary Table 4). The *F*-statistic of the two instrumental SNPs was 30.77, indicating proper strength of the instrumental variables. Multiple MR methods, including inverse variance-weighted (p = 9.46E-03, OR=1.272), maximum likelihood (p = 2.77E-12, OR=1.311), and inverse variance-weighted methods for multiplicative random effects (p = 9.46E-03, OR=1.272), showed significant associations between schizophrenia and the corresponding instrumental SNPs (Supplementary Table 5). As heterogeneity of the instrumental SNPs was detected (*p* < .05), the result of the inverse variance weighted method for multiplicative random effects was finally adopted for proper evaluation of the detected causal effect (OR = 1.272, 95%CI: 1.061–1.525, *p* = 9.46E-3; Table 2). Forest and funnel plots of single SNP effect estimates are shown in Figure 1D.

**Table 2.** Results of the bidirectional two-sample Mendelian randomization analyses.

| Exposure | Outcome | IV SNPs (n) | MR methods | Beta | SE | OR (95%CI) | P value |
| --- | --- | --- | --- | --- | --- | --- | --- |
| Niacin response | Schizophrenia | 2 | Inverse variance weighted (multiplicative random effects) | 0.240 | 0.093 | 1.272 (1.061 to 1.525) | **9.46E-03** |
| Schizophrenia | Niacin response | 19 | MR Egger | -0.274 | 0.998 | 0.760 (0.108 to 5.373) | 0.787 |
|  |  |  | Weighted Median | 0.036 | 0.131 | 1.037 (0.801 to 1.342) | 0.783 |
|  |  |  | Inverse Variance Weighted | 0.050 | 0.099 | 1.051 (0.866 to 1.275) | 0.615 |
|  |  |  | Simple Mode | 0.043 | 0.217 | 1.043 (0.681 to 1.598) | 0.847 |
|  |  |  | Weighted Mode | 0.043 | 0.232 | 1.043 (0.662 to 1.646) | 0.857 |

Note: IV, instrumental variable; SNP, single nucleotide polymorphism; MR, Mendelian randomization. Statistically significant P value has been indicated in bold.

### 3.3 The causal effect of schizophrenia on niacin response

We conducted an MR estimation using 19 instrument SNPs (Supplementary Table 4) of schizophrenia on niacin responses to assess the reverse causal effect. The result revealed none of the MR methods supported a positive causal effect of schizophrenia on the niacin response (all *p* > .05; Table 2). Heterogeneity in the dataset was not detected (*p* > .05), and no genetic pleiotropic effects were observed (*p* > .05). Scatter and forest plots of the single SNP effects are shown in Figure 1E.

## 4. Discussion

The attenuated response to niacin in patients with schizophrenia has been observed in multiple countries and populations, including the UK^10,11^, Poland^12^, America^13^, Canada^14^, Germany^15^, Netherlands^16^, Sweden^17^, China Taipei^18^ and mainland China^5^. Our results suggest that an attenuated niacin response may cause schizophrenia but not vice versa. Previous longitudinal studies have suggested a potential causal effect between the two events in terms of the sequence of occurrence, that is, niacin bluntness has been observed in patients with a clinically high risk of psychosis ^19^. Moreover, those who ultimately developed psychosis (mainly schizophrenia) were found to show less erythema (blood flow) in niacin flushing tests than those who did not at the baseline, indicating that niacin bluntness preceded the onset of psychosis and was associated with disease onset ^20^. In addition, individuals with schizophrenia and niacin bluntness have more severe negative symptoms^21^ and cognitive impairments^22^ compared to patients with schizophrenia without niacin bluntness. As we know, cognitive impairment are important for predicting psychosis ^23^. However, these studies only provide partial evidence for the potential causation, while this study offered direct confirmation of the causal effect of niacin bluntness on schizophrenia.

The arachidonic acid-GPR109A-prostaglandin pathway is a widely accepted biological mechanism underlying niacin-induced skin flushing ^24^. GPR109A and arachidonic acid are functional molecules located on the cell membrane. Several studies have explored the membrane fatty acids to identify the molecular basis of niacin bluntness. Patients with schizophrenia who failed to flush with niacin showed significantly reduced levels of arachidonic acid and docosahexaenoic acid in their red blood cell membranes^10^. Our team has also conducted a comprehensive lipidomic study of red blood cell membranes in individuals with schizophrenia and found that excessive oxidized lipids accumulated on membranes^25^. Further, their membrane fluidity was significantly affected, implying a diminished capacity of transmembrane signal transduction in these patients. In addition, a previous in vivo MRS study suggested that those with schizophrenia and poor phospholipid-related signal transduction might have increased cerebral energy metabolism, leading to obsessive oxidative stress production ^26^. We propose oxidative stress as an upstream pathogenic element since it is quite destructive, especially towards membrane lipids. A growing body of evidence supports its pivotal role in the early phases of schizophrenia ^27^. Moreover, inflammation may accompany this process ^20^. Further studies are needed to delineate the role of reactive oxygen species in the development of schizophrenia, especially the onset of the disease among the niacin-blunted subgroup ^28,29^.

This study had several limitations. First, the sample size of the cohort for GWAS of the niacin response was relatively small. However, the instrumental variables in this study were selected following standard criteria, guaranteeing their eligibility and conferring reliability on our conclusions. Moreover, this cohort has its own advantages. First, we only recruited patients and not normal individuals, as the distribution of niacin response scores in normal individuals seriously skews towards the flushing (not-blunted) end. A more dispersed distribution of the target phenotype (i.e., flushing scores in this study) only exists among patients with schizophrenia and is necessary for an efficient GWAS. Second, using a cohort with the same disease background can eliminate potential confounders and help find true associated SNPs for niacin response. Therefore, in this preliminary GWAS, we used a limited but high-quality cohort of samples, which guaranteed meaningful results.

Further, we used the total score of the 12 flushing subscores in the niacin skin test to indicate niacin response, as the total score is the most representative variable that could reflect both the amount and speed of flushing. Future studies should explore either aspect of the niacin response using other indicators (e.g., EC_50_ for speed or Sub-score_max_ for amount). Third, heterogeneity was detected between the two instrumental SNPs for niacin response in this study, potentially weakening our MR results. However, we conducted an inverse variance-weighted method, especially for multiplicative random effects, to deal with the heterogeneity, and the forest plot (Figure 1D) exhibited a robust total causal effect integrated from these two SNPs.

In conclusion, our results suggest that an attenuated niacin response causes schizophrenia but not vice versa, making relevant etiological research reasonable and promising for a future biomarker-guided personalized treatment. Further studies with larger sample sizes are warranted to validate this finding, and additional etiological studies on this distinctive niacin-blunted subgroup of schizophrenia patients are needed.

## Data Availability

All data produced in the present study are available upon reasonable request to the authors.

