## Supplementary figures and images for "Causal associations between niacin bluntness and schizophrenia: a GWAS and Mendelian randomization study"

### supplementary figure

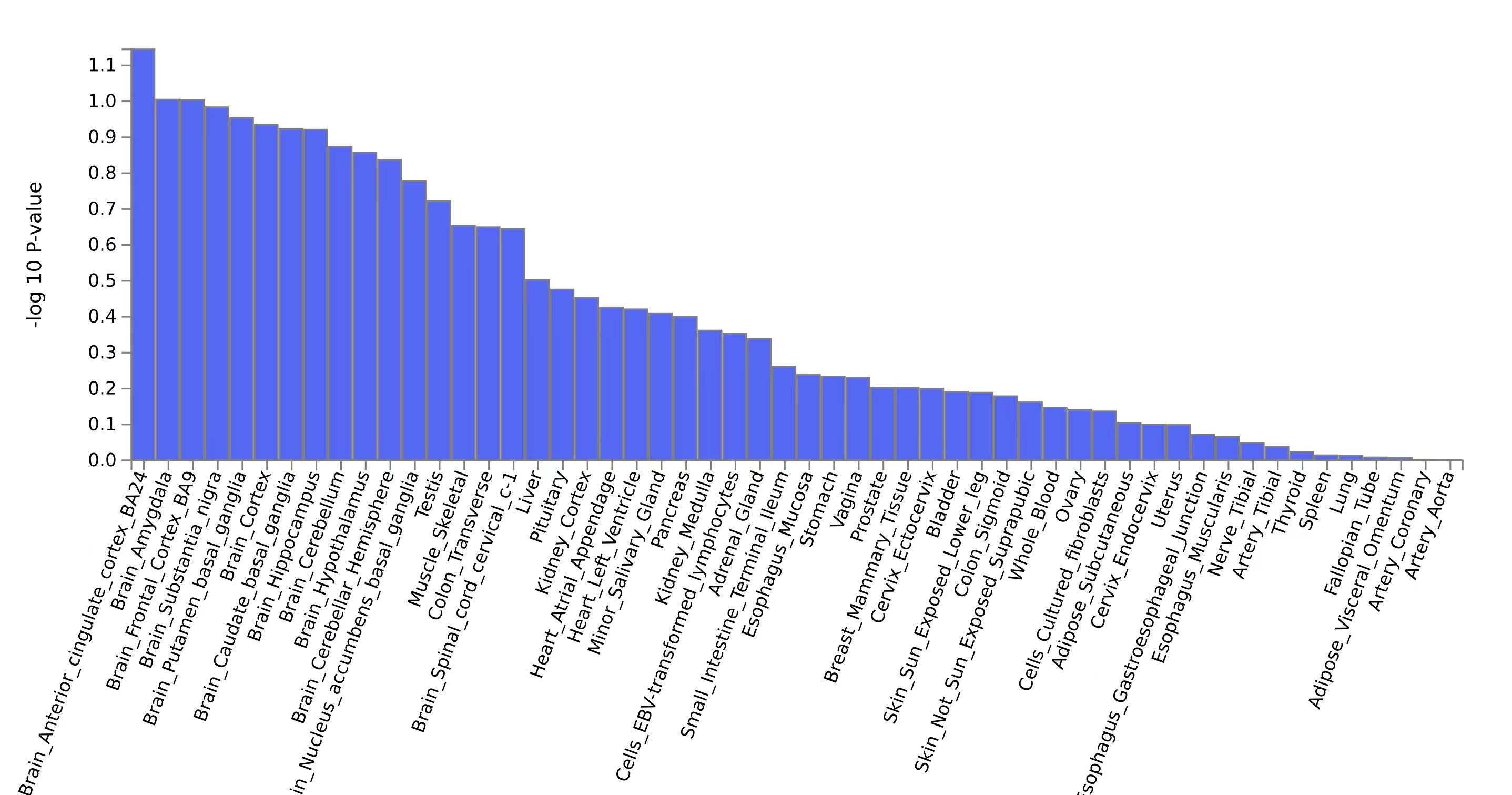
